# Low-cost predictive models of dementia risk using machine learning and exposome predictors

**DOI:** 10.1101/2023.05.03.23289444

**Authors:** Marina Camacho, Angélica Atehortúa, Tim Wilkinson, Polyxeni Gkontra, Karim Lekadir

## Abstract

Diagnosing dementia, a syndrome that currently affects more than 55 million people worldwide, remains a particularly challenging and costly task. It may involve undertaking several medical tests such as brain scans, cognitive tests and genetic tests to determine the presence and degree of cognitive decline. These procedures are associated with long procedures, subjective evaluations and high costs. As a result, patients are often diagnosed at a late stage, when symptoms become highly pronounced. Therefore, there is an urgent need for developing new strategies for early, accurate and cost-effective dementia screening and risk prediction. To overcome current limitations, we explored readily available exposome predictors for identifying individuals at risk of dementia and compared traditional statistical modeling and advanced machine learning.

From approximately 500,000 participants from the UK Biobank, 1523 participants diagnosed with dementia after their baseline assessment visit were included in our study. An equal number of healthy participants were randomly selected as the control group by matching statistical age mean and sex distribution. This resulted in a total of 3046 participants being selected for our study; 2740 participants from 19 of the 22 UK Biobank assessment centers were used for internal validation, and 306 participants from the remaining three centers were selected for external validation. We include data from the participants’ baseline visit and selected 128 low-cost exposome factors related to life course exposures that may be easily acquired through simple questionnaires. Subsequently, data imputation was performed to account for missing patient data. Two different predictive models were assessed for discriminating between participants that remained healthy and participants diagnosed with dementia after the baseline visit, i.e. (1) a classical logistic regression linear classifier and (2) a machine learning ensemble classifier based on XGBoost. We interpreted the results by estimating feature importance within the predictive models.

Our results demonstrate that machine learning models based on exposome data can reliably identify individuals that will be diagnosed with dementia. The XGBoost based model outperforms logistic regression model, achieving a mean AUC of 0.88 in the external validation tests. We identified novel exposome factors that might be combined as potential markers for dementia, such as facial aging, the frequency of use of sun/ultraviolet light protection, and the length of mobile phone use. Finally, we propose a novel neurocognitive assessment test that could be used as an online tool to screen individuals at risk of dementia for enrolment in preventive interventions and future clinical trials.

## Introduction

Diagnosing dementia, a syndrome that currently affects more than 55 million people worldwide, remains a particularly challenging task ^1,2^. It involves undertaking several exams such as cognitive tests (*e*.*g*. Montreal assessment), genetic tests (*e*.*g*. APOE) or brain scans to determine the degree of cognitive decline^3–6^. These procedures are associated with subjective evaluations, equipment related dependencies, elevated costs ^7–10^, as well as ethical concerns, such as socioeconomic and racial inequalities that can delay disease diagnosis^11^. As a result, patients are often diagnosed at a late stage when symptoms become highly pronounced^12^, while treatments do not yet exist for dementia^13–15^. Therefore, in recent years, there has been a growing focus on early diagnosis, risk prediction and preventive interventions by means of reduction of modifiable risk factors^16,17^. To this end, new low-cost, accessible and objective screening tools for identifying individuals at risk of dementia are essential.

Several predictive models for dementia risk have been proposed in the literature. For example, Li et al. proposed a method for risk prediction based on brain imaging data^18^. However, brain imaging is an expensive exam that is usually not available at the risk prediction stage. Tai et al. proposed a predictive model using biological predictors including genetic risk and cardiometabolic multimorbidity index, but their models also require the acquisition and collection of expensive biological data in the clinics^19^. Reinke et al. published a predictive model based on machine learning, which used clinical information extracted from healthcare reports as predictors, such as medication, medical history and biomarkers, hence it is unsuitable for cost-effective self-assessment of dementia risk based on easily accessible predictors. Recently, Licher et al. externally evaluated four prediction models for dementia risk that use socio-demographic and clinical predictors but found the models to be comparable to using age alone as a predictor of dementia^20^.

The use of exposome data, which provide a comprehensive description of the lifelong exposure history of an individual, has not been extensively explored and might enable the development of an accurate screening tool, and may provide insights into the underlying mechanisms behind diseases that cause dementia. In this context, a significant amount of research has focused on assessing the link between certain exposures and dementia, or diseases that might cause dementia using machine learning (ML) models, such as logistic regression^21–28^. Exposome features are of particular interest as they are not only cost effective for risk prediction, but they may be associated with beneficial lifestyle changes^23^, even though further research is needed to determine the causal relationships between the exposome and dementia^29^. In addition, several studies have reported that environmental exposures have a huge effect on pathologies due to the impact they have on cells by inducing epigenetic changes^30–32^.

In this paper, we present a novel ML model to identify individuals at risk of dementia based on low-cost exposome predictors. We compare its performance to a traditional model based on statistical methods and report the most important exposome factors relevant for dementia prediction. We used the UK Biobank data and performed nested cross-validation, and an external validation in three different assessment centers^33,34^. Finally, we propose a novel assessment test that could be used as a tool to screen individuals at risk for dementia for enrolment in future clinical trials and preventive interventions.

## Methods

An overview of the study design is provided in Figure 1. In brief, first, we selected 128 readily available exposome features from the UK Biobank. Subsequently, we performed data pre-processing, including selection of the study population, data cleaning and imputation. We then trained and evaluated our ML models using internal and external validation cohorts. Last but not least, we studied the most important features for the model decision to interpret the results and identify potential novel markers. Each step of our pipeline is described in more detail in the following subsections.

**Fig. 1.**
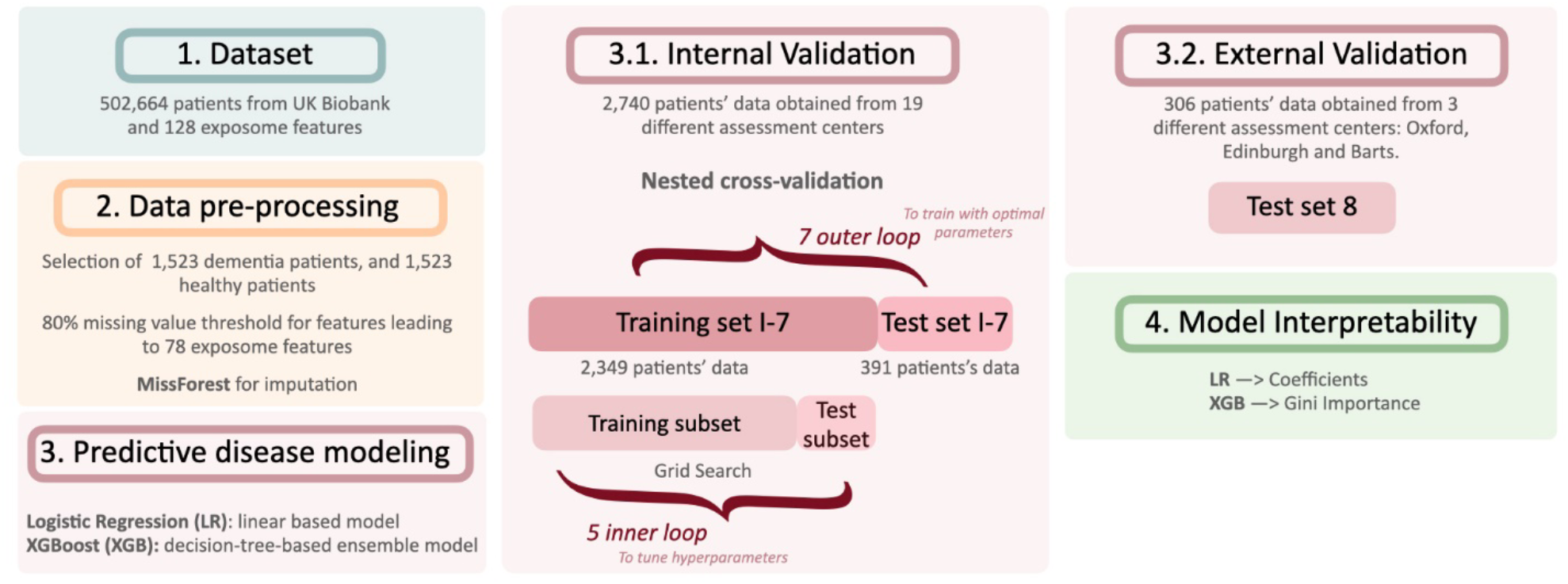
Overview of our methodology for identifying individuals at risk of dementia using exposome data from the UK Biobank.

### 1. Dataset

This work has been conducted using the UK Biobank resource under application reference number 65769. UK Biobank (UKBB) is a large-scale, population-based cohort of half a million participants, who were aged 40 to 69 years when recruited from across the United Kingdom between 2006 and 2010^33,34^. The cohort data is established as an open access resource for public research and can be access under request. At recruitment, a wide variety of phenotypic information and biological samples were collected for the participants.

The UK Biobank study was approved by the National Information Governance Board for Health and Social Care and the National Health Service North West Multicentre Research Ethics Committee. All participants provided signed informed consent at enrolment and all research was performed in accordance with relevant guidelines/regulations. All data used in this analysis is available through application to the UK Biobank.

### 2. Data pre-processing

#### Dementia diagnosis

Dementia cases were defined using the International Standard Classification of Diseases and Related Health Problems, 10^th^ edition (ICD-10) diagnostic codes related to dementia disease as primary or secondary diagnosis at the time of this analysis^35^. These codes are provided in the Supplementary Table 1.

#### Study population

We selected individuals who developed dementia at least one year after the last baseline assessment, to ensure a wash-out period of at least one year. Specifically, while the baseline data collection took place during 2006-2010, only individuals with a dementia diagnosis after 2011 were selected, while participants with a diagnosis of dementia before 2011 were excluded. This resulted in a (total of 1,523 individuals included in the study. Additionally, we selected a control group consisting of 1,523 healthy individuals with two constraints: (i) both sexes were equally represented in the dementia and control groups, and (ii) the mean age of the healthy group was the same as the dementia group. Note that we decided not to exclude deceased individuals in order not to create a bias in our machine learning model against individuals at higher risk of mortality.

#### Exposome features

For the scope of this study, a group of 128 exposome factors was selected from the UK Biobank from six main categories: traumatic events, sociodemographic, physical measurements, lifestyle, environmental factors and early life factors. These factors are life course exposures collected during the first assessment visit (baseline assessment). We excluded features with more than 20% of missing data, resulting in a total of 78 features included in the study. A detailed list of those 78 exposome variables used in this work is provided in Supplementary Table 2.

#### Data imputation

We imputed missing values using the MissForest algorithm. The method is based on random forests trained on the observed values to predict missing data^36^. We applied the imputation separately for the dementia patients and the control population to reduce the time complexity, as well as for the internal and external validation datasets respectively. We implemented it using the missForest R package version 1.4 and the algorithm’s default parameters^37^.

### 3. Predictive modeling of dementia

We used a traditional statistical learning-based model, Logistic Regression (LR), and a state-of-the art machine learning model based on parallel tree boosting, XGBoost (XGB). The LR and XGB models were implemented in Python 3.7.4 using the Scikit-Learn library 0.24.2^38^. A detailed list of the hyper-parameters values under consideration is provided in Supplementary Table 3.

We performed nested cross-validation (CV) with 7 outer and 5 inner folds using the data from the 19 assessment centers. The inner cross-validation folds were used for hyperparameter tuning by means of grid-search, while the outer for internal validation.

#### Model interpretability

One of the added values of this work is the identification of the exposome attributes with the highest impact in the models’ decision on whether to classify individuals at risk of dementia or not. These features are of particular interest as they could be potential novel markers in screening dementia. The list of the most important features involved in the LR models’ decision is obtained by the coefficients of the model. The list of the most relevant features involved in the XGB models’ decision is obtained by means of the state-of-the-art Gini importance, also known as mean decrease impurity. Gini importance is computed by subtracting the sum of squares probabilities of each class from one, which corresponds to the likelihood of a given variable being wrongly classified when selected randomly^39^.

Additionally, we included the interpretation of our best models by Shapley Additive exPlanations (SHAP) as ML is known as a “black-box” due to difficulties in explaining the way of extracting the results. This methodology is suitable for the explanation of complex models, such as XGB, providing feature importance values as well as the direction of contribution of each feature to the final prediction^40^.

#### Models’ performance analysis

We evaluated the model performance in the internal and external validation cohorts in terms of sensitivity (ratio of correct positive classifications), precision (positive predictive value), F1 score (harmonic mean between precision and sensitivity), and AUC (probability of ranking a randomly chosen positive instance higher than a randomly chosen negative one). We conducted internal and external validation; a detailed explanation is described as follows: i) in internal validation, a nested cross-validation was performed using data from 19 of 22 UK Biobank assessment centres with 90% of dementia cases including 2, 740 participants ii) in external validation, a validation was performed using data from 3 of 22 UK Biobank assessment centres (Oxford, Edinburgh and Barts) with 10% of dementia cases including 306 participants.

For our study, we are particularly interested in obtaining both accurate detection of patients that will result in being diagnosed with the disorder (high sensitivity), and accurate detection of those without that condition (high specificity). For this reason, we used AUC as the most relevant statistic for our analysis as it describes the relationship between the true positive classifications and the true negative classifications, meaning it measures the overall performance considering the specificity and sensitivity. Moreover, the classification power of an algorithm can be evaluated accordingly to the AUC value that it achieves: no discrimination [AUC=0.5], poor discrimination [0.6≥AUC>0.5], acceptable discrimination [0.7≥AUC>0.6], excellent discrimination [0.8≥AUC>0.7], or outstanding discrimination [AUC>0.9]^41^.

#### Experimental design

To analyze the predictive power of the exposome features, we used a varying set of features. More precisely, to ensure that the models’ predictive power was not solely related to the inclusion of age, a factor known to play an important role in previous predictive dementia models^20^, we compared the models’ performance when using the following set of features: (i) all available exposome data including age (78 features), (ii) all available exposome data without including age (77 features), (iii) age alone.

Subsequently, we selected the 30 variables with the highest Gini importance values for the best model, *i*.*e*. the XGB model based on 78 exposome features including age. We subsequently rejected the variables that we considered expensive to acquire or difficult to measure, such as the average daytime level of noise. As previously, we repeated the comparison with the varying set of predictive factors, i.e., exposome data including age (30 features) and exposome data without age (30 features).

## Results

Table 1 provide the mean and standard deviation of the evaluation metrics (AUC, F1-score, Precision, Sensitivity) in the internal and external validation cohorts using logistic regression and XGBoost.

**Table 1.**
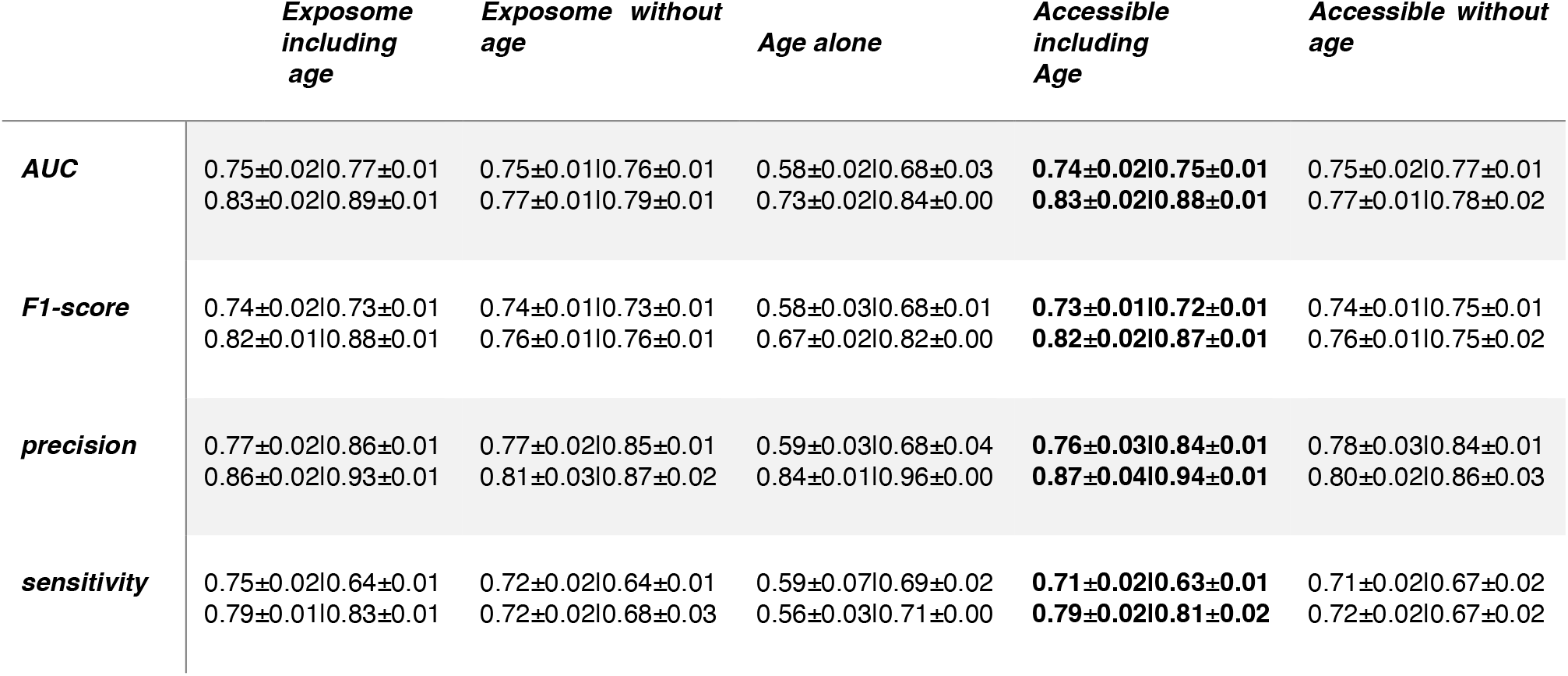
Top and bottom row of each metric corresponds to logistic regression and XGBoost performance, based on varying sets of features; (i) 78 exposome features including age, (ii) 77 exposome features without considering age, (iii) age, (iv) 30 easily accessible exposome features including age, and (v) 30 easily accessible features without considering age. Mean and standard deviation of each performance metric is provided, for the internal and external validation cohorts.

Despite the popularity of LR for identifying patients at risk, in all experimental settings, the more advanced machine learning algorithm, XGB, outperformed LR^20^. More precisely, in internal validation, applying XGB instead of LR provides an increase in the mean AUC on all the models; +0.079 for the model based on 78 exposome variables including age, +0.020 for the model based on the 77 exposome variables excluding age, and +0.089 in the model based solely on age. In external validation, the AUC improvement achieved by XGB is +0.120, +0.030, and +0.130, for the models based on all exposome features, on all exposome features except age, and on age alone, respectively.

Accessible exposome based models performed similarly to those using all exposome features. In internal validation, the use of accessible features instead of the complete list of exposome data resulted in a mean AUC decrease of -0.010 by using LR and age. Nevertheless, by using the LR model based on easily accessible exposome features except age or when using XGB as the modelling approach, no decrease in accuracy was observed. Furthermore, in the external validation, the use of accessible features without age implied a mean AUC improvement of +0.010 by using LR, and for the rest of the cases, a mean AUC loss of -0.010. Hence, the models based on a reduced set of exposome features have still an excellent discrimination with the best model being the one with accessible variables including age, which achieved a mean AUC equal to 0.83 in internal validation, and 0.88 in external validation. Additionally, precision was higher, meanwhile sensitivity was lower.

Figures 2 and 3 allow for a visual comparison between the LR and XGB models. The XGB performance (as a purple line) outperforms the LR (as a green line) in all the exposome-based experiments and metrics. Moreover, a detailed description of the importance of the models’ variables if provided in Supplementary Material Tables 4-11.

**Fig. 2.**
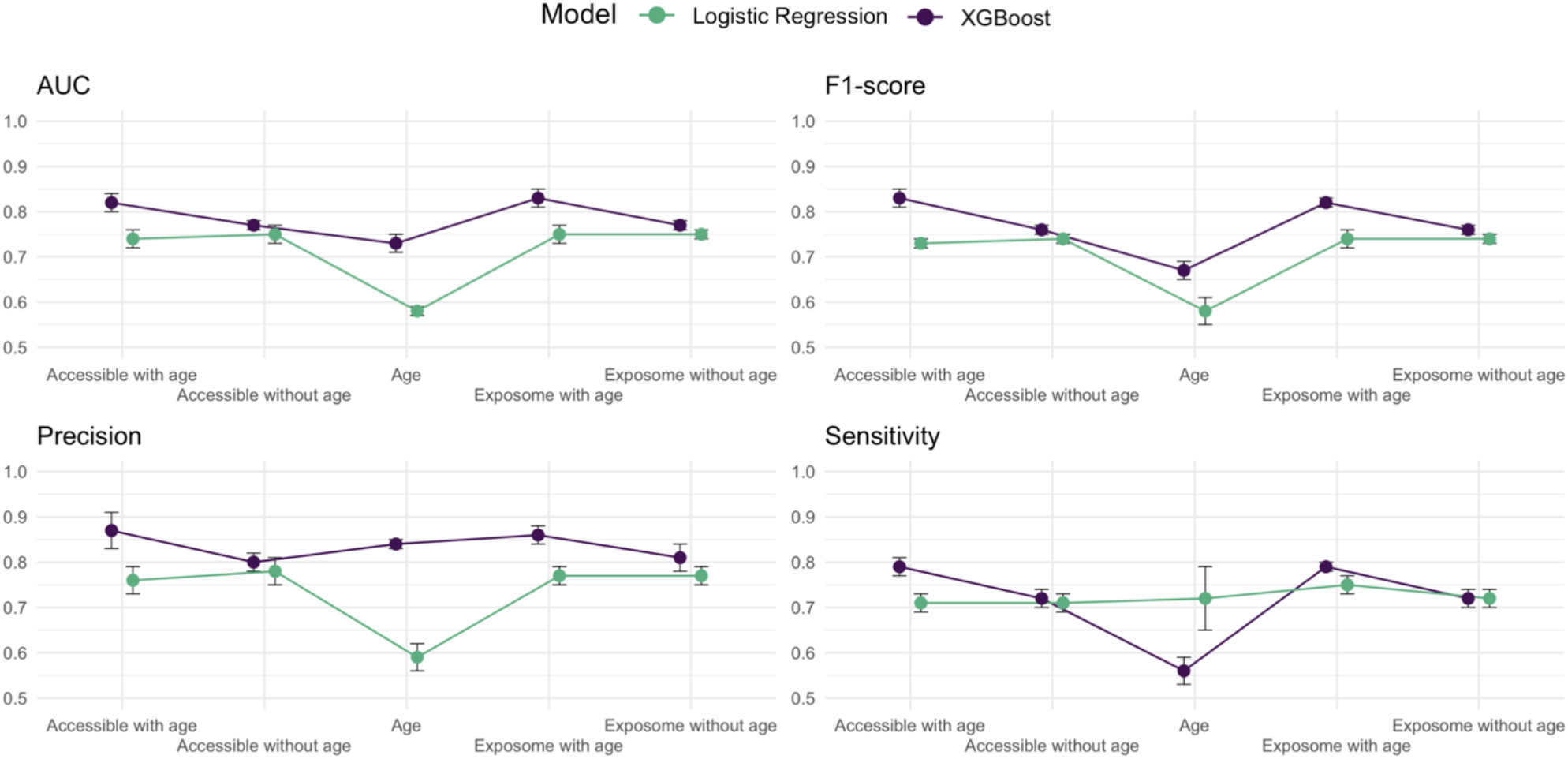
Internal Validation results for logistic regression and XGBoost performance in terms of AUC, F1-score, precision and sensitivity. Logistic regression and XGBoost results are represented with green and dark purple, respectively.

**Fig. 3.**
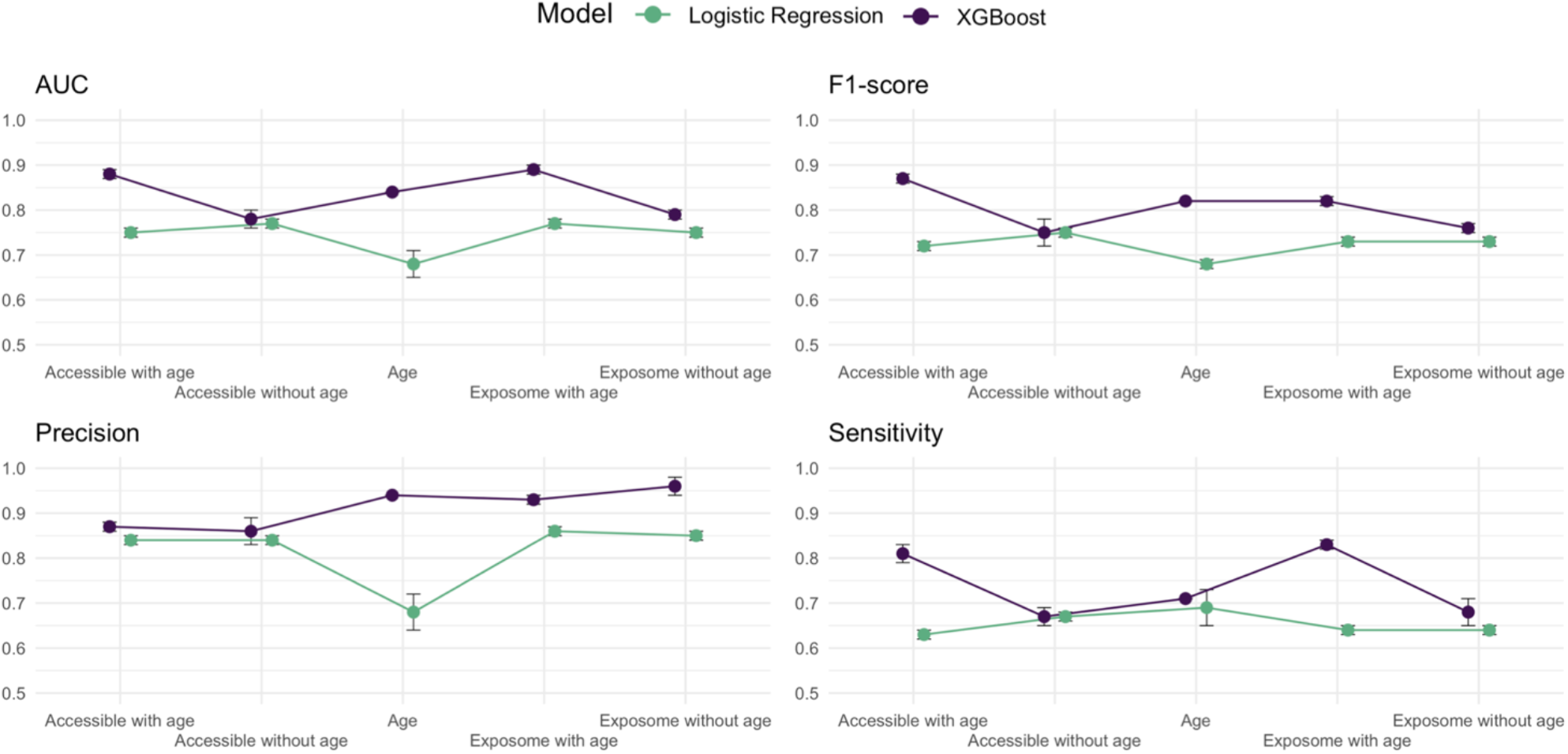
External Validation results for logistic regression and XGBoost performance in terms of AUC, F1-score, precision and sensitivity. Logistic regression and XGBoost results are represented with green and dark purple, respectively.

Overall, the model that provided the best balance between performance and number of input variables required was the XGB model based on 30 exposome features including age. Most of the relevant variables in the LR and XGB models were related to education, diet, and sleep. Interestingly, other factors that were highly correlated with dementia were facial ageing, the frequency of use of sun/ultraviolet light protection, and the length of mobile phone use. Feature importance for the thirty exposome features that are common to the LR and XGboost models is presented in Figures 4 and 5. To assess feature importance as a relative measure in LR, we use standardised coefficients based on the approach proposed by Menard^42^. It should be noted that the ordering of the predictors would be the same regardless of the choice of method for standardizing the LR coefficients ^43^.

**Fig. 4.**
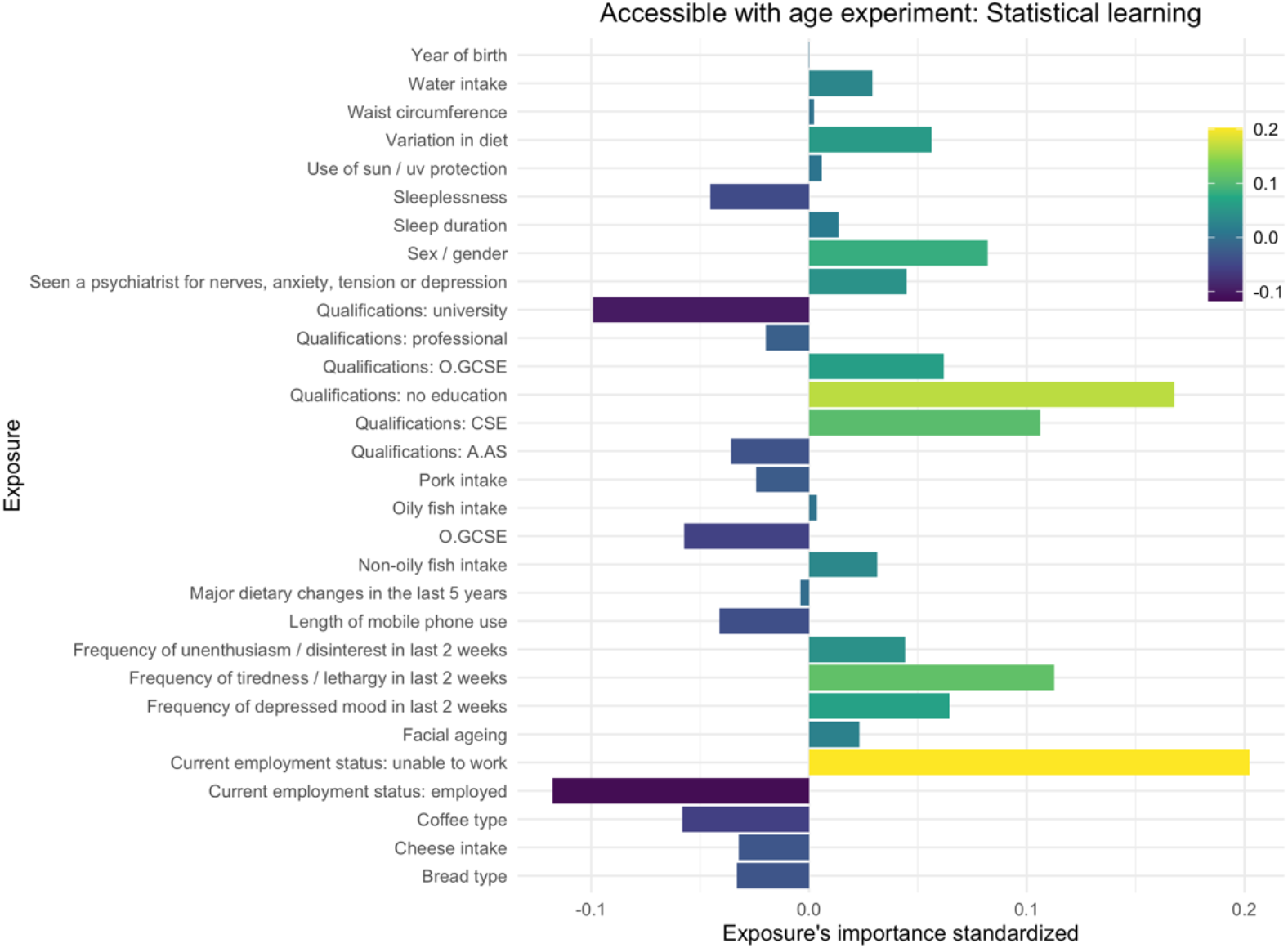
Importance of exposome factors in logistic regression model decision. Precisely, thirty accessible exposome variables including age and their importance based on the StandardizedCoefficient for Logistic Regression Formula.

**Fig. 5.**
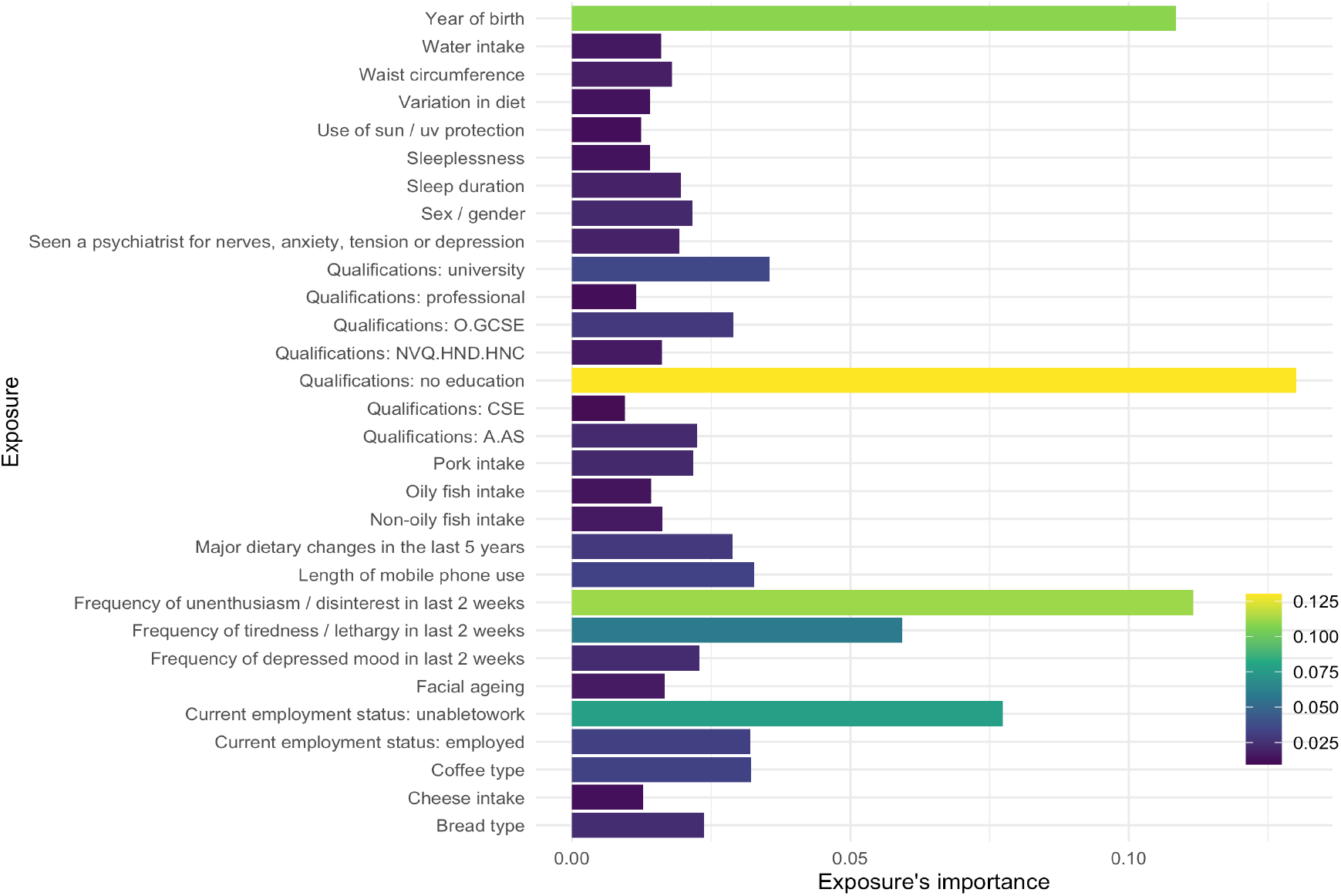
Importance of exposome factors in XGBoost model decision. Precisely, thirty accessible exposome variables including age and their importance based on the mean decrease importance are provided.

Furthermore, we report feature importance by Shapley Additive exPlanations (SHAP)-values to identify the most important features in a more robust way and, additionally, assess the direction of the feature effect for the best models^44,45^. The results are provided in figures 6 and 7.

**Fig. 6.**
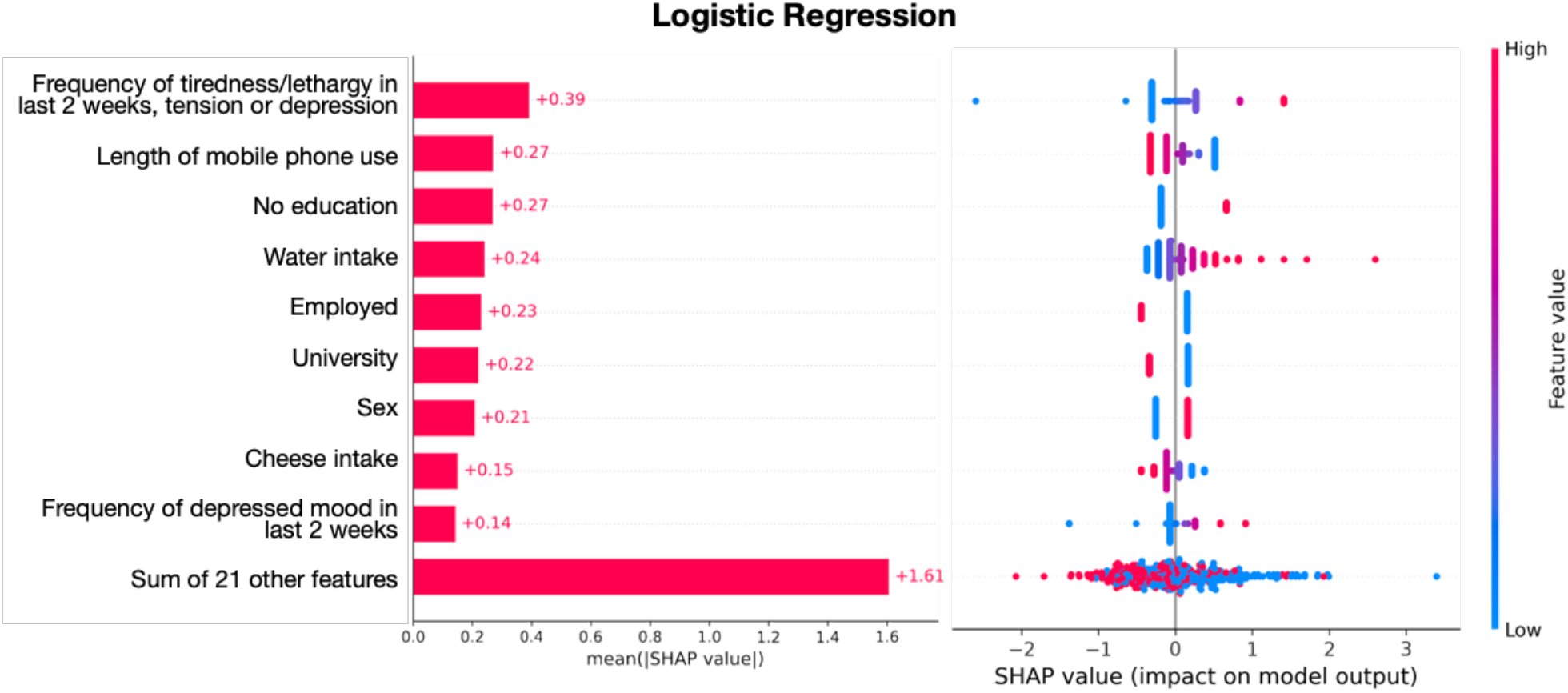
Logistic Regression explainability. On the left side, the importance of each feature as expressed by the mean absolute SHAP value is provided. On the right side, the impact of each feature on the model output is shown. Higher values of the features are indicated by red, while lower with blue.

**Fig. 7.**
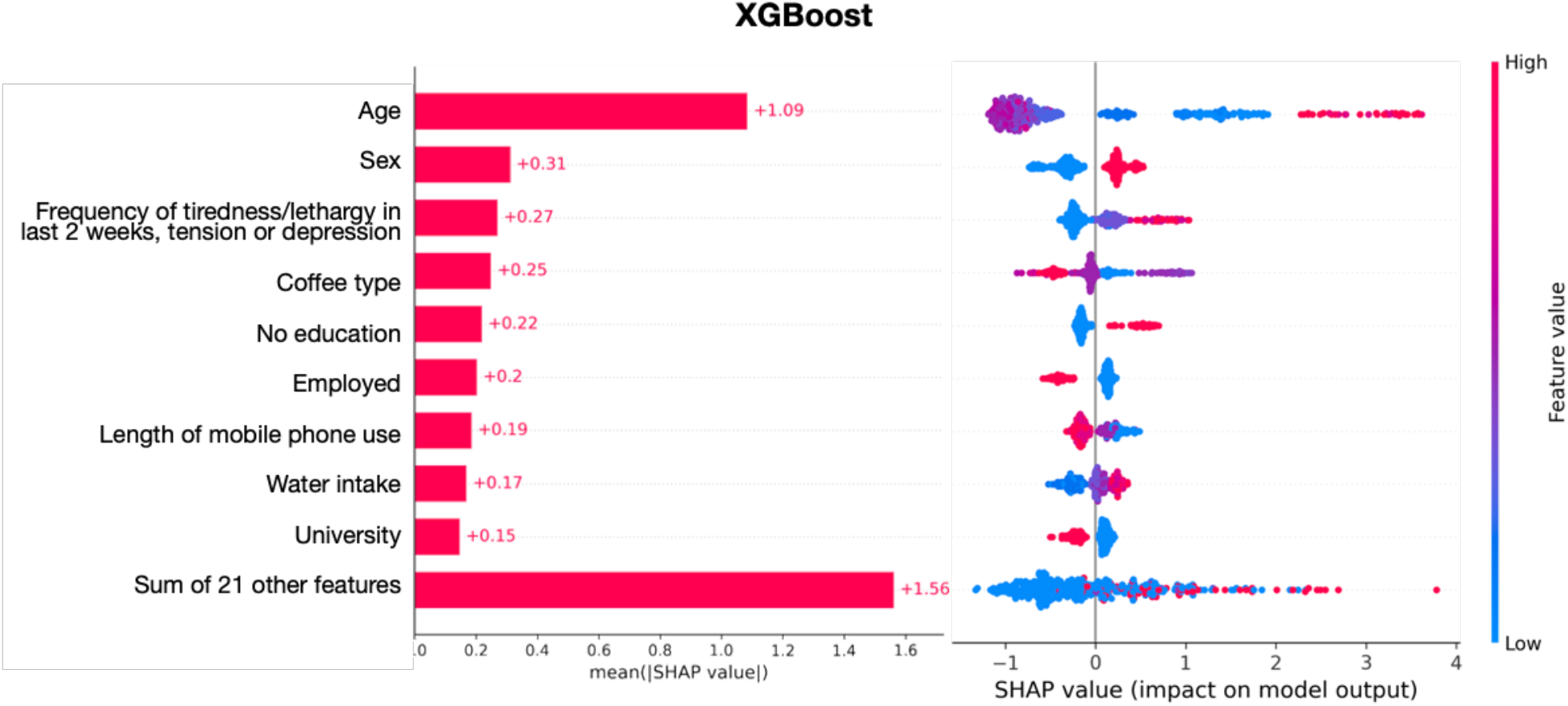
XGBoost explainability. On the left side, the importance of each feature as expressed by the mean absolute SHAP value is provided. On the right side, the impact of each feature on the model output is shown. Higher values of the features are indicated by red, while lower with blue.

Moreover, LR and XGB models attributed similar importance to most of the variables. Nevertheless, some variables had much more importance in XGB. Not having received any type of major education, apathy, and the type of consumed bread were more relevant in the XGB model. Also, the year of birth had no relevance in LR. However, it was one of the most important variables in the XGB. This may be due to the capacity XGB has to uncover high dimensional relationships between age and the other variables for an individual to end up being at risk of dementia.

## Discussion and Conclusions

This exploratory study aimed to identify individuals at risk of being diagnosed with dementia by exploring the most relevant low-cost and easily acquirable exposome features. Even though we are only using exposome predictors and no biological and clinical predictors, we achieve competitive performance during internal evaluation tests, compared to existing works^20,46^. In particular, for external validation, we obtained a 0.88 AUC value, compared to an AUC of 0.49-0.92 in the recent work by Licher et al.^20^, whose predictive models included clinical predictors as well. These comparisons must be interpreted with care, as the studies have used different cohorts and populations, and in this work, the external validation tool place in the same country albeit in different assessment centres. However, our results provide indications regarding the added value of exposome predictors for dementia risk prediction, which can be easily and inexpensively collected through self-assessment questionnaires.

XGB based on decision trees outperformed LR. This is most probably related to the capacity of XGB to capture multidimensional and complex relations among the heterogeneous exposome data. Furthermore, variables related to education, sleep, diet, and patient’s feelings were the most relevant in all our models, which is consistent with current literature, and highlights the importance of individuals’ exposure effects through life^47–49^. However, such studies did not have a holistic approach, they focused on a specific exposome feature or subgroup of them and used only statistical models.

We were able to discover new exposure predictors of dementia risk, which have received limited attention in the past, such as the use of sun/UV protection and face aging. These results are aligned with some of the existing research studies on these topics. For instance, Gao *et al*. revealed a positive association between dementia and time spent in the sun^50^, which in turn may be related to the use of sun/UV protection. Furthermore, a recent artificial intelligence model was able to discriminate faces of individuals with and without dementia ^51^. One possible hypothesis is that one of the face characteristics that allow this discrimination may be related to an older appearance of the individuals, which our variable “face aging” might be capturing. However, these are hypotheses that would require further research to uncover the precise relationships and causality between these exposures and dementia. Another direction for future research identified by our study involves the interplay between sleeplessness, sleep duration and dementia. The results of our study add to previous works that pointed out that the associations of sleep and dementia are yet to be clarified^52–54^.

Considering that ML models are at risk of overfitting, we applied several techniques to reduce this risk of poor performance in other settings. We used a nested cross-validation and multiple imputations to prevent a biased model performance as a result of deletion or single imputation of participants’ missing data^55^. More precisely, we used an advanced method for imputation, the MissForest algorithm, to ensure accuracy and computational efficiency, as it can handle different types of data and outperforms other established methods, such as k-nearest neighbours imputation or multivariate imputation using chained equations^36,56^.

Another strength of this study is that our models could be easily implemented as a web-based tool, where people could fill a questionnaire to provide exposome data information and would then be provided with a report of their potential dementia risk. This tool could be used to screen individuals who might of interest for future epidemiological and drug studies, and for preventive intervention.

Despite the significance of the study, there exist some limitations. First, although, to the best of our knowledge, this is the most complete study in terms of number of exposome factors considered, we explored as potential predictors the features that were available to us through the application reference number 65769. Nonetheless, there might exist other variables that might further improve the classification accuracy. On the other hand, the final reduced models containing 30 variables have the advantage of using only easily acquired features. Additionally, the exposome data was collected by questionnaires which could introduce a reporting and recall bias.

Moreover, we used ICD-10 codes to identify participants with dementia for inclusion in this study. Even though this method has been validated against medical records, it will not be able to identify dementia cases perfectly^35^. For instance, the proportion of true dementia cases identified (sensitivity) is unknown^57^. However, we do know the positive predictive value (the likelihood a dementia case truly does have dementia) is high (80-86%) ^35^. An additional validation strategy could be applied in future works of dementia risk assessment to minimize false-negative and false-positive diagnoses. A work in this direction is that of Doblhammera et al., 2015 involving a validation approach for dementia cases to minimize false-positives in Medicare data^58^.

Limitations in terms of methodology include the imputation methodology which may be further improved by being applied it in each fold. However, we decided not to adopt this approach to avoid high computational costs as it has been previously reported not to lead to meaningful differences on performance^59^. Furthermore, the external validation step is crucial to ensure that reported metrics are not overestimated. In an effort to simulate it, we took advantage of the design of UKBB as it comprises data from 22 different assessment centers and we could test geographical generality. However, we could not externally validate them in another population above the cohort. As far as we know, there is no other cohort with similar variables to perform such validation.

Overall, this preliminary study provides consistent evidence of the exposome’s power for identifying individuals at risk of dementia when combined with advanced machine learning approaches, such as XGBoost. Our best model, based on 30 readily available and low-cost variables, could serve as a screening tool for timely patient referral for further examination, intervention and for enrolment in future clinical trials. We identified novel potential markers, such as face aging and use of sun/UV protection. Finally, our results demonstrate the potential of artificial intelligence techniques to improve and update current predictive models in healthcare.

## Data Availability

The data that support the findings of this study are available from UK Biobank, but restrictions apply to the availability of these data, which were used under license for the current study, and so are not publicly available. Data are however available from the authors upon reasonable request and with permission of UK Biobank.

## Acknowledgements

This research has received funding from the European Union’s Horizon 2020 research and innovation program under Grant Agreement Nº 848158 (EarlyCause) and Grant Agreement Nº 874739 (LongITools), and from the Spanish Ministry of Science, Innovation, and Universities within the framework of the ‘‘Retos Investigación’’ program, project RTI2018-099898-B-I00 (HeartBrainCom).

## Author Contribution

M.C, A.A and K.L had full access to all of the data and take responsibility for the integrity of the data and the accuracy of the data analysis. M.C., A.A., K.L., and T.W. designed the study. All authors contributed to the interpretation of the findings. M.C., A.A. and T.W. drafted the paper and all authors contributed to the final version.

