## Supplementary Material for "Low-cost predictive models of dementia risk using machine learning and exposome predictors"

*[1]BCN-AIM laboratory. Mathematics and Computer Science Faculty. University of Barcelona. Gran Via de les Corts Catalanes, 585. 08007 Barcelona, Spain*
*[2]Centre for Clinical Brain Sciences, The University of Edinburgh, Edinburgh, EH16 4SB, UK*
*[3]Usher Institute, The University of Edinburgh, Edinburgh, EH16 4SB, UK*

### Supplementary Material

| | ICD-10 codes |
|---|---|
| Alzheimer's disease | F000\|F001\|F002\|F009\|G300\|G301\|G308\|G309 |
| Vascular dementia | F010\|F011\|F012\|F013\|F018\|F019\|I673 |
| Frototemporal dementia | F020\|G310 |
| Other causes of dementia | A810\|F021\|F022\|F023\|F024\|F028\|F051\|F106\|G311\|G318 |
| All causes of dementia | F000\|F001\|F002\|F009\|G300\|G301\|G308\|G309\|F010\|F011\|F012\|F013\|F018\|F019\|I673\|F020\|G310\|A810\|F021\|F022\|F023\|F024\|F028\|F051\|F106\|G311\|G318 |

**Table 1. ICD-10 dementia codes list.** Used codes for selecting dementia cases of interest in UKBB for our study.

| Code | Name | Name in predictive disease modeling |
|---|---|---|
| 31 | sex/ gender | sex |
| 48 | Waist circumference | waistcircum |
| 49 | Hip circumference | hipcircum |
| 50 | Standing height | standingheight |
| 21001 | Body mass index (BMI) | bmi |
| 21002 | Weight | weight |
| 23099 | Body fat percentage | bodyfatpercent |
| 23101 | Whole body fat-free mass | wholebodyfatfreemass |
| 23102 | Whole body water mass | wholebodywatermass |
| 23105 | Basal metabolic rate | basalmetabolicrate |
| 34 | Age | age |
| 189 | Townsend deprivation index at recruitment | townsendeprivation |
| 757 | Time employed in main current job | timeinjob |
| 767 | Length of working week for main job | lengthworkweek |
| 796 | Distance between home and job workplace | distancehomework |

| | | |
|---|---|---|
| 806 | Job involves mainly walking or standing | jobwalkingstand |
| 816 | Job involves heavy manual or physical work | jobphysical |
| 826 | Job involves shift work | jobshiftwork |
| 6142 | Current employment status | Recoded as:<br>-7.0: 'otherwork',<br>1.0: 'employed',<br>2.0: 'retired',<br>3.0: 'lookingafterhome',<br>4.0: 'unabletowork',<br>5.0: 'unemployed',<br>6.0: 'unpaidwork',<br>7.0: ‘student' |
| 6138 | Qualifications | Recoded as:<br>-7.0: 'noeducation',<br>1.0: 'university',<br>2.0: 'A/AS',<br>3.0: 'O/GCSE',<br>4.0: 'CSE',<br>5.0: 'NVQ/HND/HNC',<br>6.0: 'professional' |
| 845 | Age completed full time education | agecompleted |
| 1160 | Sleep duration | sleepduration |
| 1200 | Sleeplessness | sleeplessness |
| 1220 | Daytime dozing / sleeping (narcolepsy) | daytimedozing |
| 1190 | Nap during day | napduringday |
| 1239 | Current tobacco smoking | currenttobacco |
| 1249 | Past tobacco smoking | pasttobacco |
| 20414 | Frequency of drinking alcohol | frequdrinkingalcohol |
| 20453 | Ever taken cannabis | evertakencannabis |
| 20411 | Ever been injured or injured someone else through drinking alcohol | everbeeninjuredrinkalcohol |
| 20405 | Ever had known person concerned about, or recommend reduction of, alcohol consumption | everhadknownpersonalcoh |
| 20117 | Alcohol drinker status | alcoholdrinkerstatus |
| 1050 | Time spend outdoors in summer | timespendsummer |
| 1060 | Time spent outdoors in winter | timespendswinter |
| 2267 | Use of sun/uv protection | usesunprotection |
| 1747 | Hair colour (natural, before greying) | haircolour |
| 1757 | Facial ageing | facialageing |
| 1727 | Ease of skin tanning | easeofskin |
| 1110 | Length of mobile phone use | lengthmobileuse |
| 1120 | Weekly usage of mobile phone in last 3 months | weeklyusemobile |
| 1140 | Difference in mobile phone use compared to two years previously | differencemobile2years |
| 1150 | Usual side of head for mobile phone use | usualsideofheadmobile |
| 2237 | Plays computer games | playcomputergames |
| 1289 | Cooked vegetable intake | cookedvegetable |
| 1299 | Salad / raw vegetable intake | saladintake |
| 1309 | Fresh fruit intake | fruitintake |
| 1319 | Dried fruit intake | driedfruit |

| | | |
|---|---|---|
| 1329 | Oily fish intake | oilyfishintake |
| 1339 | Non-oily fish intake | nonoilyfish |
| 1349 | Processed meat intake | processedmeat |
| 1359 | Poultry intake | poultryintake |
| 1369 | Beef intake | beefintake |
| 1379 | Lamb/mutton intake | lambintake |
| 1389 | Pork intake | porkintake |
| 1408 | Cheese intake | cheeseintake |
| 1418 | Milk type used | milkused |
| 1438 | Bread intake | breadintake |
| 1448 | Bread type | breadtype |
| 1458 | Cereal intake | cerealintake |
| 1468 | Cereal type | cerealtype |
| 1478 | Salt added to food | salttofood |
| 1488 | Tea intake | teaintake |
| 1498 | Coffee intake | coffeintake |
| 1508 | Coffee type | coffetype |
| 1548 | Variation in diet | variationdiet |
| 1428 | Spread type | spreadtype |
| 1528 | Water intake | waterintake |
| 1538 | Major dietary changes in the last 5 years | dietarychange |
| 2654 | Non-butter spread type details | nonbutterspread |
| 100240 | Coffee consumed | coffeeconsumed |
| 100390 | Tea consumed | teaconsumed |
| 100580 | Alcohol consumed | alcoholconsumed |
| 104670 | Vitamin supplement user | closettomajorroad |
| 24009 | Traffic intensity on the nearest road | trafficintensity |
| 24014 | Close to major road | closetomajorroad |
| 24020 | Average daytime sound level of noise pollution | averagedaytimenoisepoll |
| 24021 | Average evening sound level of noise pollution | averageeveningnoise |
| 1737 | Childhood sunburn occasions | childhoodsunburn |
| 1677 | Breastfed as a baby | breastfedbaby |
| 1687 | Comparative body size at age 10 | bodysizeat10 |
| 1697 | Comparative height size at age 10 | bodyheightat10 |
| 1707 | Handedness (chirality/laterality) | handedness |
| 1767 | Adopted as a child | adoptedaschild |
| 1777 | Part of a multiple birth | partmultiplebirth |
| 1787 | Maternal smoking around birth | maternalsmoking |
| 20491 | Someone to take to doctor when needed as a child | someonetakedoctorchild |
| 20490 | Sexually molested as a child | sexuallymolestedchild |

| | | |
|---|---|---|
| 20489 | Felt loved as a child | feltlovedchild |
| 20488 | Physically abused by family as a child | physicallyabusedfamiliychild |
| 20487 | Felt hated by family member as a child | felthatedfamiliychild |
| 20531 | Victim of sexual assault | victimsexualassault |
| 20530 | Witnessed sudden violent death | witnessedsuddenviolentdeath |
| 20529 | Victim of physically violent crime | victimphysicallyviolentcrime |
| 20526 | Been in serious accident believed to be life-threatening | beenseriousaccidentlifethrea |
| 20525 | Able to pay rent/mortgage as an adult | abletopaymortgageadult |
| 20524 | Sexual interference by partner or ex-partner without consent as an adult | sexualinterferenceadult |
| 20523 | Physical violence by partner or ex-partner as an adult | physicalviolenceadult |
| 20522 | Been in a confiding relationship as an adult | beenconfidingrelationadult |
| 20521 | Belittlement by partner or ex-partner as an adult | belittlementbypartneradult |
| 20498 | Felt very upset when reminded of stressful experience in past month | upsetstressfulpastmonth |
| 20497 | Repeated disturbing thoughts of stressful experience in past month | repeatedisturbingpastmonth |
| 20495 | Avoided activities or situations because of previous stressful experience in past month | avoidstresspasthmonth |
| 2040 | Risk taking | risktaking |
| 2050 | Frequency of depressed mood in last 2 weeks | freqdepressed |
| 2090 | Seen doctor (GP) for nerves, anxiety, tension or depression | seendoctordepress |
| 2100 | Seen a psychiatrist for nerves, anxiety, tension or depression | seenpsychiatrist |
| 4526 | Happiness | happiness |
| 4598 | Ever depressed for a whole week | everdepressed |
| 4609 | Longest period of depression | perioddepress |
| 4620 | Number of depression episodes | numberdepressper |
| 20126 | Bipolar and major depression status | bipolarstatus |
| 20127 | Neuroticism score | neuroticismscore |
| 4631 | Ever unenthusiastic/disinterested for a whole week | everdesinteresedweek |
| 2080 | Frequency of tiredness / lethargy in last 2 weeks | freqtiredness2weeks |
| 2070 | Frequency of tenseness / restlessness in last 2 weeks | freqtensenness2weeks |
| 2060 | Frequency of unenthusiasm / disinterest in last 2 wee | frequnenthusiasm2weeks |

**Table 2. Exposome variables.** Features codes, brief description, and name used in the predictive disease modeling in this study. For further details see: https://biobank.ndph.ox.ac.uk/ukb/search.cgi.

| | |
|---|---|
| **Logistic Regression (LR)** | LR's solver, penalty and c value parameters were selected from values: ['newton-cg', 'lbfgs','liblinear'], ['l2','l1'], [100, 10, 1.0,0.1,0.01], respectively. |
| **XGBoost (XGB)** | XGB's learning rate, minimum child weight, gamma value, subsample, subsampling of columns by tree and maximum depth parameters were selected from values: [0.05, 0.10, 0.15, 0.20, 0.25, 0.30], [1, 5, 10], [0.5, 1, 5], [0.6, 0.8, 1.0], [0.6, 0.8, 1.0], [3, 4, 5], respectively. |

**Table 3. Models parameters.** Set parameters to select the best one by grid search for logistic regression and XGBoost.

| Feature names | Feature importance |
|---|---|
| unabletowork | 0.7990948707932082 |
| noeducation | 0.7701467689536782 |
| freqtiredness2weeks | 0.5521478748730597 |
| CSE | 0.49221176844154985 |
| bodyheightat10 | 0.3475859589741969 |
| sex | 0.32816656973739244 |
| freqdepressed | 0.30528118966086604 |
| NVQ.HND.HNC | 0.29736678060358546 |
| unemployed | 0.2876308020502356 |
| variationdiet | 0.2579342290485306 |
| adoptedaschild | 0.22903796478707344 |
| frequnenthusiasm2weeks | 0.20555365774448695 |
| seendoctordepress | 0.19267447511842398 |
| partmultiplebirth | 0.19172287487473572 |
| daytimedozing | 0.16478234500003536 |
| nonoilyfish | 0.16368224011515625 |
| waterintake | 0.14064069847193966 |
| processedmeat | 0.12974476429792248 |
| seenpsychiatrist | 0.1258084157967755 |
| lookingafterhome | 0.12459112006359803 |
| facialageing | 0.11059786563296763 |
| sleepduration | 0.0857789710497489 |
| otherwork | 0.07402433191959656 |
| salttofood | 0.07037096742733877 |
| usesunprotection | 0.06992997328065328 |
| townsendeprivation | 0.06825911155910416 |
| handedness | 0.049238421024302946 |
| waistcircum | 0.04793899338601537 |
| oilyfishintake | 0.04756499312935988 |
| beefintake | 0.041327865816626896 |

| | |
|---|---|
| teaintake | 0.03842100361093446 |
| saladintake | 0.03158845793858962 |
| hair | 0.029925786573811168 |
| driedfruit | 0.022219423637495662 |
| fruitintake | 0.0190311602440867 |
| easeofskin | 0.018454610343071762 |
| averagedaytimenoisepoll | 0.009545346012323729 |
| spreadtype | 0.009541043442048553 |
| cookedvegetable | 0.006597657831816788 |
| maternalsmoking | 0.003348408581254331 |
| age | 0.0028901709762333507 |
| milkused | 0.002361034991885523 |
| basalmetabolicrate | 0.0005763735278743984 |
| trafficintensity | 7.248033444484082e-06 |
| wholebodywatermass | -0.0002541126284101278 |
| napduringday | -0.00036189623403922496 |
| student | -0.0011577063081640567 |
| averageeveningnoise | -0.005219959318777563 |
| breadintake | -0.008582767744800114 |
| bodyfatpercent | -0.00863433179796285 |
| lambintake | -0.009861463495064822 |
| hipcircum | -0.011301822599221535 |
| cerealintake | -0.012300355126245048 |
| coffeintake | -0.0155045271939493 |
| cerealtype | -0.019457266016959834 |
| dietarychange | -0.021418474719163402 |
| bmi | -0.024362361250370505 |
| freqtensenness2weeks | -0.04942681169536718 |
| weight | -0.051057228883513744 |
| unpaidwork | -0.056843825504880936 |
| standingheight | -0.057035752952442315 |
| professional | -0.06379318882016219 |
| risktaking | -0.06728433102740988 |
| playscomputergames | -0.10170800063063289 |
| bodysizeat10 | -0.1178952047338215 |
| porkintake | -0.13753935862490196 |
| poultryintake | -0.13761976879065477 |
| breadtype | -0.13963289304597626 |
| cheeseintake | -0.1725192239471509 |
| A.AS | -0.18093007365586883 |

| | |
|---|---|
| lengthmobileuse | -0.18568516826109024 |
| sleeplessness | -0.23392018856448996 |
| retired | -0.26163805008480184 |
| O.GCSE | -0.26668988179100417 |
| coffetype | -0.3039836107872154 |
| university | -0.4784998715390053 |
| employed | -0.8170492843174261 |

**Table 4. Logistic Regression experiment i importance.** Beta coefficients of the best model in experiment i, including all 78 exposome features, and age.

| Feature names | Feature importance |
|---|---|
| frequnenthusiasm2weeks | 0.09683714 |
| noeducation | 0.08710915 |
| age | 0.059220415 |
| unabletowork | 0.034068488 |
| freqtiredness2weeks | 0.029432375 |
| university | 0.022348212 |
| lengthmobileuse | 0.022141721 |
| employed | 0.021175394 |
| coffetype | 0.0203274 |
| breadtype | 0.018557763 |
| sex | 0.017414859 |
| O.GCSE | 0.01607513 |
| dietarychange | 0.015276172 |
| waterintake | 0.014869789 |
| facialageing | 0.013350801 |
| A.AS | 0.013197278 |
| sleepduration | 0.01317547 |
| sleeplessness | 0.012937212 |
| seenpsychiatrist | 0.01283299 |
| cheeseintake | 0.012107989 |
| townsendeprivation | 0.01209624 |
| porkintake | 0.012067366 |
| maternalsmoking | 0.011710942 |
| basalmetabolicrate | 0.011669567 |
| cerealtype | 0.011595745 |
| beefintake | 0.011581679 |
| wholebodywatermass | 0.011460858 |

| | |
|---|---|
| processedmeat | 0.011313159 |
| seendoctordepress | 0.011286292 |
| freqdepressed | 0.01125522 |
| bodysizeat10 | 0.011059561 |
| bmi | 0.010976969 |
| variationdiet | 0.010882125 |
| waistcircum | 0.010794337 |
| bodyfatpercent | 0.010625474 |
| partmultiplebirth | 0.010562446 |
| oilyfishintake | 0.010523526 |
| freqtensenness2weeks | 0.010363069 |
| averageeveningnoise | 0.010293973 |
| standingheight | 0.010266351 |
| cookedvegetable | 0.010188464 |
| retired | 0.010187101 |
| hair | 0.009992661 |
| saladintake | 0.009669156 |
| driedfruit | 0.009631438 |
| breadintake | 0.009503574 |
| coffeintake | 0.009432134 |
| usesunprotection | 0.009430321 |
| easeofskin | 0.009429669 |
| averagedaytimenoisepoll | 0.009406129 |
| hipcircum | 0.00939616 |
| bodyheightat10 | 0.009382897 |
| weight | 0.009209668 |
| nonoilyfish | 0.008945363 |
| milkused | 0.008925849 |
| daytimedozing | 0.008819742 |
| risktaking | 0.008750045 |
| poultryintake | 0.00869977 |
| teaintake | 0.008587085 |
| CSE | 0.0077471174 |
| lambintake | 0.007619098 |
| spreadtype | 0.007522983 |
| cerealintake | 0.007461837 |
| playscomputergames | 0.006775147 |
| trafficintensity | 0.0055928617 |
| professional | 0.0048531927 |
| unemployed | 0.0 |

| | |
|---|---|
| adoptedaschild | 0.0 |
| lookingafterhome | 0.0 |
| handedness | 0.0 |
| unpaidwork | 0.0 |
| student | 0.0 |
| otherwork | 0.0 |
| salttofood | 0.0 |
| napduringday | 0.0 |
| fruitintake | 0.0 |
| NVQ.HND.HNC | 0.0 |

**Table 5. XGBoost experiment i importance.** Mean decrease in impurity of the best model in experiment i, including all 78 exposome features, and age.

| Feature names | Feature importance |
|---|---|
| unabletowork | 0.8221937926546908 |
| noeducation | 0.7675864941346321 |
| freqtiredness2weeks | 0.5534205112262925 |
| CSE | 0.5016681903303138 |
| bodyheightat10 | 0.3412490326701457 |
| unemployed | 0.31095545155387644 |
| sex | 0.30828367545531016 |
| freqdepressed | 0.3046012738379921 |
| NVQ.HND.HNC | 0.2935526453611111 |
| variationdiet | 0.2623041834417304 |
| adoptedaschild | 0.24558175262836618 |
| frequnenthusiasm2weeks | 0.20588458303593807 |
| partmultiplebirth | 0.19655918517164728 |
| seendoctordepress | 0.19041704681144037 |
| nonoilyfish | 0.16417932917808378 |
| daytimedozing | 0.16206956948845397 |
| waterintake | 0.13967328734917506 |
| processedmeat | 0.13006003430489393 |
| seenpsychiatrist | 0.12415174033602341 |
| lookingafterhome | 0.12345128210723871 |
| facialageing | 0.11113640295952834 |
| sleepduration | 0.08720852659662283 |
| otherwork | 0.07902847421399335 |
| salttofood | 0.07242955228038964 |
| usesunprotection | 0.07070171699333955 |

| | |
|---|---|
| bmi | 0.0697949272242189 |
| townsendeprivation | 0.06824872734439949 |
| handedness | 0.05245931343820933 |
| waistcircum | 0.04869985350920041 |
| oilyfishintake | 0.04789328165690254 |
| beefintake | 0.043397940445142155 |
| teaintake | 0.03911727861599759 |
| averagedaytimenoisepoll | 0.032537786457409276 |
| saladintake | 0.03135458972912431 |
| hair | 0.03052879099844987 |
| driedfruit | 0.02323386035376982 |
| fruitintake | 0.020191558113989018 |
| easeofskin | 0.018018421051116618 |
| spreadtype | 0.009788947821495341 |
| cookedvegetable | 0.007406849384847426 |
| maternalsmoking | 0.002652035929515488 |
| milkused | 0.0022287887700811154 |
| basalmetabolicrate | 0.001018090077374355 |
| trafficintensity | 6.812061661808984e-06 |
| napduringday | -0.00025326055928498806 |
| student | -0.0020445521556046392 |
| bodyfatpercent | -0.008555262858277462 |
| breadintake | -0.0088148612298773 |
| hipcircum | -0.011628264710005809 |
| lambintake | -0.012517021976832514 |
| cerealintake | -0.012683401372007042 |
| coffeintake | -0.013838208681440247 |
| cerealtype | -0.016501090810151425 |
| dietarychange | -0.02103206017896381 |
| standingheight | -0.024902423703724294 |
| averageeveningnoise | -0.027172976437131357 |
| freqtensenness2weeks | -0.04761616371002392 |
| unpaidwork | -0.05848843022902942 |
| wholebodywatermass | -0.059242381212978874 |
| professional | -0.06286811399582126 |
| risktaking | -0.06601016562802531 |
| weight | -0.09249603095292466 |
| playscomputergames | -0.09950447067559445 |
| bodysizeat10 | -0.11030199730689244 |
| poultryintake | -0.13576281223878608 |

| | |
|---|---|
| porkintake | -0.13777841191811416 |
| breadtype | -0.14040572296529955 |
| cheeseintake | -0.1745681315780744 |
| A.AS | -0.17616888575950002 |
| lengthmobileuse | -0.18517464768416828 |
| sleeplessness | -0.22825162840087304 |
| retired | -0.2519438036832187 |
| O.GCSE | -0.2701652567286506 |
| coffetype | -0.3038404139291029 |
| university | -0.48679020466164813 |
| employed | -0.8018142025060747 |

**Table 6. Logistic regression experiment ii importance.** Beta coefficients of the best model in experiment ii, including all 78 exposome features without age.

| Feature names | Feature importance |
|---|---|
| noeducation | 0.097557396 |
| frequnenthusiasm2weeks | 0.07145809 |
| freqtiredness2weeks | 0.03995111 |
| unabletowork | 0.037726734 |
| university | 0.030256988 |
| freqdepressed | 0.021143224 |
| coffetype | 0.02084257 |
| lengthmobileuse | 0.019693406 |
| employed | 0.01835606 |
| sex | 0.017739411 |
| dietarychange | 0.016949574 |
| seenpsychiatrist | 0.016367147 |
| nonoilyfish | 0.014502648 |
| A.AS | 0.013606643 |
| NVQ.HND.HNC | 0.0135592325 |
| professional | 0.013492333 |
| oilyfishintake | 0.013481283 |
| breadtype | 0.01338525 |
| waistcircum | 0.013164152 |
| CSE | 0.012783184 |
| variationdiet | 0.012663714 |
| waterintake | 0.012498501 |
| porkintake | 0.012203903 |
| partmultiplebirth | 0.012181109 |

| | |
|---|---|
| townsendeprivation | 0.012173437 |
| sleepduration | 0.01208716 |
| sleeplessness | 0.011940707 |
| O.GCSE | 0.011854045 |
| cheeseintake | 0.011847415 |
| facialageing | 0.011473552 |
| wholebodywatermass | 0.011159522 |
| usesunprotection | 0.010899673 |
| standingheight | 0.01073008 |
| seendoctordepress | 0.0107283285 |
| processedmeat | 0.010727502 |
| lambintake | 0.010651219 |
| saladintake | 0.0106444815 |
| trafficintensity | 0.010411065 |
| playscomputergames | 0.010391578 |
| basalmetabolicrate | 0.010343207 |
| bmi | 0.010072267 |
| unpaidwork | 0.0100471815 |
| cookedvegetable | 0.010023229 |
| easeofskin | 0.009842227 |
| risktaking | 0.009795964 |
| bodysizeat10 | 0.009444613 |
| teaintake | 0.009437719 |
| driedfruit | 0.009157308 |
| breadintake | 0.009000501 |
| averagedaytimenoisepoll | 0.008950245 |
| cerealtype | 0.008818506 |
| maternalsmoking | 0.0088102715 |
| hipcircum | 0.008806865 |
| fruitintake | 0.008571381 |
| milkused | 0.008538478 |
| averageeveningnoise | 0.008507117 |
| bodyfatpercent | 0.008324287 |
| spreadtype | 0.008275652 |
| hair | 0.008243092 |
| bodyheightat10 | 0.0077080885 |
| cerealintake | 0.0076690675 |
| freqtensenness2weeks | 0.0074736443 |
| salttofood | 0.00736396 |
| weight | 0.006995288 |

| | |
|---|---|
| coffeintake | 0.0067807827 |
| daytimedozing | 0.0065324395 |
| poultryintake | 0.0063453442 |
| beefintake | 0.0061339578 |
| retired | 0.006129549 |
| lookingafterhome | 0.0061034923 |
| handedness | 0.0053618993 |
| napduringday | 0.0031088975 |
| adoptedaschild | 0.0 |
| otherwork | 0.0 |
| unemployed | 0.0 |
| student | 0.0 |

**Table 7. XGBoost experiment ii importance.** Mean decrease in impurity of the best model in experiment ii, including all 78 exposome features without age.

| Feature names | Feature importance |
|---|---|
| age | 0.19712915967827074 |
| waistcircum | 0.06652347789494986 |
| freqtiredness2weeks | 0.06175376129280421 |
| coffetype | 0.057829357472101085 |
| noeducation | 0.051950244801691706 |
| frequnenthusiasm2weeks | 0.04958490408061094 |
| lengthmobileuse | 0.04199666393338608 |
| waterintake | 0.03941187641590733 |
| sleepduration | 0.033372389752630606 |
| cheeseintake | 0.03057612366838708 |
| freqdepressed | 0.029528293472942647 |
| university | 0.02611444161538413 |
| oilyfishintake | 0.026035063032909766 |
| breadtype | 0.023888704170135264 |
| usesunprotection | 0.02312466223149328 |
| porkintake | 0.022232961032675747 |
| facialageing | 0.02162576980684671 |
| dietarychange | 0.019741336674152225 |
| nonoilyfish | 0.019589290944922445 |
| employed | 0.01935632446122216 |
| unabletowork | 0.019297604940834497 |
| O.GCSE | 0.017948681243271754 |
| variationdiet | 0.01766634898435299 |

| | |
|---|---|
| sleeplessness | 0.01752267758823033 |
| A.AS | 0.01586395897271834 |
| professional | 0.012726091599974582 |
| sex | 0.012393369549484862 |
| seenpsychiatrist | 0.012090643669719684 |
| NVQ.HND.HNC | 0.008734821982219842 |
| CSE | 0.004390995035769205 |

**Table 8. Logistic regression experiment iv importance.** Beta coefficients of the best model in experiment iv, including 30 exposome features with age.

| Feature names | Feature importance |
|---|---|
| noeducation | 0.13005702 |
| frequnenthusiasm2weeks | 0.111596994 |
| age | 0.10847546 |
| unabletowork | 0.07737419 |
| freqtiredness2weeks | 0.059287462 |
| university | 0.03547091 |
| lengthmobileuse | 0.03277825 |
| coffetype | 0.032158088 |
| employed | 0.03201079 |
| O.GCSE | 0.028986074 |
| dietarychange | 0.028874753 |
| breadtype | 0.023724632 |
| freqdepressed | 0.02286569 |
| A.AS | 0.022486519 |
| porkintake | 0.021780878 |
| sex | 0.021703193 |
| sleepduration | 0.019509457 |
| seenpsychiatrist | 0.019267462 |
| waistcircum | 0.017962595 |
| facialageing | 0.01662292 |
| nonoilyfish | 0.016265951 |
| NVQ.HND.HNC | 0.016185502 |
| waterintake | 0.015985902 |
| oilyfishintake | 0.01421486 |
| variationdiet | 0.014067943 |
| sleeplessness | 0.01403257 |
| cheeseintake | 0.012774249 |
| usesunprotection | 0.012388791 |

| | |
|---|---|
| professional | 0.011532689 |
| CSE | 0.009558272 |

**Table 9. XGBoost experiment iv importance.** Mean decrease in impurity of the best model in experiment iv, including 30 exposome features with age.

| Feature names | Feature importance |
|---|---|
| unabletowork | 6.347584215684726 |
| noeducation | 1.0430940064601182 |
| CSE | 0.9377385693861648 |
| sex | 0.9162772096211732 |
| freqtiredness2weeks | 0.5909167024914921 |
| NVQ.HND.HNC | 0.3858928362401906 |
| freqdepressed | 0.3649140615759508 |
| variationdiet | 0.3091653029058656 |
| seenpsychiatrist | 0.2309381530497048 |
| frequnenthusiasm2weeks | 0.2118858076508404 |
| nonoilyfish | 0.17005792816577237 |
| waterintake | 0.153352228287531 |
| facialageing | 0.11964617161388968 |
| sleepduration | 0.07177104851284946 |
| usesunprotection | 0.044449009375211554 |
| oilyfishintake | 0.02972911021180139 |
| waistcircum | 0.01204957655776865 |
| professional | -0.01672973951959676 |
| dietarychange | -0.032842099322184695 |
| standingheight | -0.033761516218602086 |
| porkintake | -0.14234661040889357 |
| A.AS | -0.14600849948940237 |
| breadtype | -0.1560646423714222 |
| cheeseintake | -0.1595462020426008 |
| lengthmobileuse | -0.21087301273978962 |
| O.GCSE | -0.2686666209039234 |
| sleeplessness | -0.2741836036151164 |
| coffetype | -0.287540570913639 |
| university | -0.4809617141454088 |
| employed | -0.6665732783669833 |

**Table 10. Logistic regression experiment v importance.** Beta coefficients of the best model in experiment v, including 30 exposome features without age.

| Feature names | Feature importance |
|---|---|
| noeducation | 0.16328414 |
| frequnenthusiasm2weeks | 0.10102846 |
| unabletowork | 0.07493863 |
| freqtiredness2weeks | 0.061514735 |
| university | 0.05077663 |
| freqdepressed | 0.03392722 |
| A.AS | 0.032755006 |
| coffetype | 0.032333333 |
| lengthmobileuse | 0.030847454 |
| seenpsychiatrist | 0.027733112 |
| employed | 0.027199315 |
| O.GCSE | 0.02534901 |
| dietarychange | 0.02409403 |
| sex | 0.022740059 |
| CSE | 0.021034457 |
| NVQ.HND.HNC | 0.020908946 |
| waterintake | 0.020471176 |
| sleepduration | 0.020035649 |
| facialageing | 0.019867834 |
| sleeplessness | 0.019399641 |
| cheeseintake | 0.01883636 |
| professional | 0.018637689 |
| porkintake | 0.018631848 |
| variationdiet | 0.018625967 |
| nonoilyfish | 0.017192842 |
| waistcircum | 0.016740926 |
| usesunprotection | 0.015747584 |
| breadtype | 0.015697617 |
| oilyfishintake | 0.015450928 |
| standingheight | 0.0141993845 |

**Table 11. XGBoost experiment v importance.** Mean decrease in impurity of the best model in experiment v, including 30 exposome features without age.
